# Biofunctionalized Two-dimensional MoS_2_ Receptors for Rapid Response Modular Electronic SARS-CoV-2 and Influenza A Antigen Sensors

**DOI:** 10.1101/2020.11.17.20233569

**Authors:** C. Muratore, M.K. Muratore, D.R. Austin, P. Look, A.K. Benton, L.K. Beagle, M.J. Motala, D.C. Moore, M.C. Brothers, S.S. Kim, K. Krupa, T.A. Back, J.T. Grant, N. R. Glavin

**Affiliations:** Department of Chemical and Materials Engineering, University of Dayton, Dayton OH 45469 USA; m-nanotech Ltd. Dayton, OH 45409, USA; Department of Biology, University of Dayton, Dayton, OH 45469 USA; UES Inc. Dayton, OH 454352 USA; Materials and Manufacturing Directorate, Air Force Research Laboratory, Wright-Patterson Air Force Base, OH 45433 USA; Department of Mechanical Engineering, Dayton, OH 45469 USA; 711^th^ Human Performance Wing, Air Force Research Laboratory, Wright-Patterson Air Force Base, OH 45433 USA; Surface Analysis Consultant, Clearwater, FL, 33767 USA

**Keywords:** COVID-19, SARS-Cov-2, Coronavirus, virus diagnostics, antigen sensors, electronic sensors, 2D materials, MoS_2_

## Abstract

Multiplex electronic antigen sensors for detection of SARS-Cov-2 spike glycoproteins or hemagglutinin from Influenza A in liquid samples with characteristics resembling extracted saliva were fabricated using scalable processes with potential for economical mass-production. The sensors utilize the sensitivity and surface chemistry of a two-dimensional MoS_2_ transducer for attachment of antibody fragments in a conformation favorable for antigen binding. Ultra-thin layers (3 nm) of amorphous MoS_2_ were directly sputtered over the entire sensor chip at room temperature and laser annealed to create an array of semiconducting 2H-MoS_2_ active sensor regions between metal contacts. The semiconducting region was functionalized with monoclonal antibody Fab (fragment antigen binding) fragments derived from whole antibodies complementary to either SARS-CoV-2 S1 spike protein or Influenza A hemagglutinin using a papain digestion to cleave the antibodies at the disulfide hinges. The high affinity for the MoS_2_ transducer surface with some density of sulfur vacancies for the antibody fragment base promoted chemisorption with antigen binding regions oriented for interaction with the sample. The angiostatin converting enzyme 2 (ACE2) receptor protein for the SARS-CoV-2 spike glycoprotein, was tethered to a hexa-histidine (his_6_) tag at its c-terminus both for purification purposes, as well as a motif for binding to MoS_2_. This modified protein was also investigated as a bio-recognition element. Electrical resistance measurements of sensors functionalized with antibody fragments and exposed to antigen concentrations ranging from 2-20,000 picograms per milliliter revealed selective responses in the presence of complementary antigens with sensitivity to SARS-CoV-2 or influenza A on the order of pg/mL and comparable to gold-standard diagnostics such as Polymerase Chain Reaction (PCR) analysis. Lack of antigen sensitivity for the larger ACE2 BRE further demonstrates the utility of the engineered antibody fragment/transducer interface in bringing the target antigen closer to the transducer surface for sensitivity required for early detection viral diagnostics.

---

Inexpensive, daily-use tests for early and rapid detection of pathogens contained within easily accessible secretions such as saliva will be critical for the safe return of individuals to work, school, and recreational/cultural activities in the midst of a pandemic. The most reliable approaches for viral analysis such as Polymerase Chain Reaction (PCR) and enzyme-linked immunosorbent assay (ELISA) provide detection of low antigen concentrations (10^−12^ g/mL), but currently require extensive sample handling, costly laboratory infrastructure, and test-to-result times ranging between hours to days due to the backlog of samples. Advances are leading to broader point-of-care capability involving these testing methods, but even this effort is insufficient for curbing the impact of the current COVID-19 pandemic and potential future global health emergencies. A survey of sensor modalities for virus diagnostics points to electronic sensors, specifically those exhibiting responses in electrical conductivity upon viral adsorption, as the most likely path toward realization of this objective of frequent, home-based testing due to their superlative sensitivity (i.e., single virus detection)^1^, ease of use, instantaneous results, and potentially low cost^2-6^. While electronic biosensor devices were first developed five decades ago^7^, only recent developments in nanomaterials have given rise to electronic sensors with sensitivity comparable that of conventional institutional diagnostics such as PCR and ELISA^1, 8-14^.

While sensitivity is well-documented for nanomaterial-based electronic sensors, selectivity remains a challenge. Reports of antibody and antibody Fab (Fragment antigen binding) fragment functionalization of electronic sensor transducers indicate their utility in terms of providing specific binding sites for analytes in solution. An early demonstration of real-time detection of single Influenza A antigen proteins was achieved through single crystal silicon nanowire transistors yielding immediate, binding-induced conductance changes^1^. These sensors were functionalized for specific detection with monoclonal antibodies immobilized by linkers to the nanowires’ native surface oxide. Erlanger et al. found that IgG antibodies adhering to fullerenes also had the flexibility to adhere to the hydrophobic, π electron-rich surface of single walled carbon nanotubes despite their sharp radius of curvature^15^. These one-dimensional materials performed well as biosensors, but were difficult to integrate into devices. The understanding developed for nanotube functionalization was applicable in some cases to two-dimensional graphene. This atomically thick material was easier to integrate into devices and demonstrated low levels of thermal and measurement noise^16^, although the small band gap resulted in responses of lower magnitude for adsorption events. Recently, antibody functionalized graphene was integrated into electronic field effect transistor-based sensors demonstrating fg/mL sensitivity for SARS-CoV-2 and excellent selectivity, however many complex and energy-intensive steps were necessary for device fabrication^17^, thereby reducing the utility of such a device for inexpensive mass production. Two-dimensional (2D) transition metal dichalcogenides (TMDs) are a recent addition to the family of low-dimensional materials possessing advantages of optical transparency and mechanical flexibility shared with graphene, but with a much larger room temperature bandgap (for monolayer MoS_2_ about 1.8 eV^18^ compared to ∼ 0.1 eV ^19^ for graphene.). The large bandgap of 2D MoS_2_ yields a lower value of subthreshold swing for FET devices^20^, where changes in charge carrier density are more sensitive to single binding events. Sarkar et al. show how the difference in band gap results in a nearly 10^2^ fold increase in sensitivity for MoS_2_ over graphene^20^.

The composition and structure of 2D TMDs enable interactions with both metal layers and chalcogen layers comprising TMD compounds. The chalcogen species are especially intriguing for antibody and antibody fragment functionalization. Figures 1A-C show how papain antibody digestion breaks the antibody at the disulfide hinge. It is expected that the now-available cysteine will possess a high affinity for a sulfur surface (with some sulfur vacancies) of the MoS_2_ transducer, promoting binding of the fragment base to the transducer and leaving the active region of the antibody available for binding. Not only does antibody digestion have the potential to promote the desired BRE conformation, but also reduces transducer-antigen distance, thereby circumventing potential issues with binding and electrostatic changes occurring outside of the Debye length. Lee et al. showed antibody functionalization of MoS_2_ with whole antibodies for prostate specific antigen^21^. More recently, reports of prostate specific antigen sensing via ‘chemically reduced’ PSA antibodies bound to thick (1 μm) MoS_2_ transducers through disulfide linkages between the MoS_2_ nanosheets and the chemically reduced antibodies with exposed sulfydryl (-SH) groups^22^.

**Figure 1:**
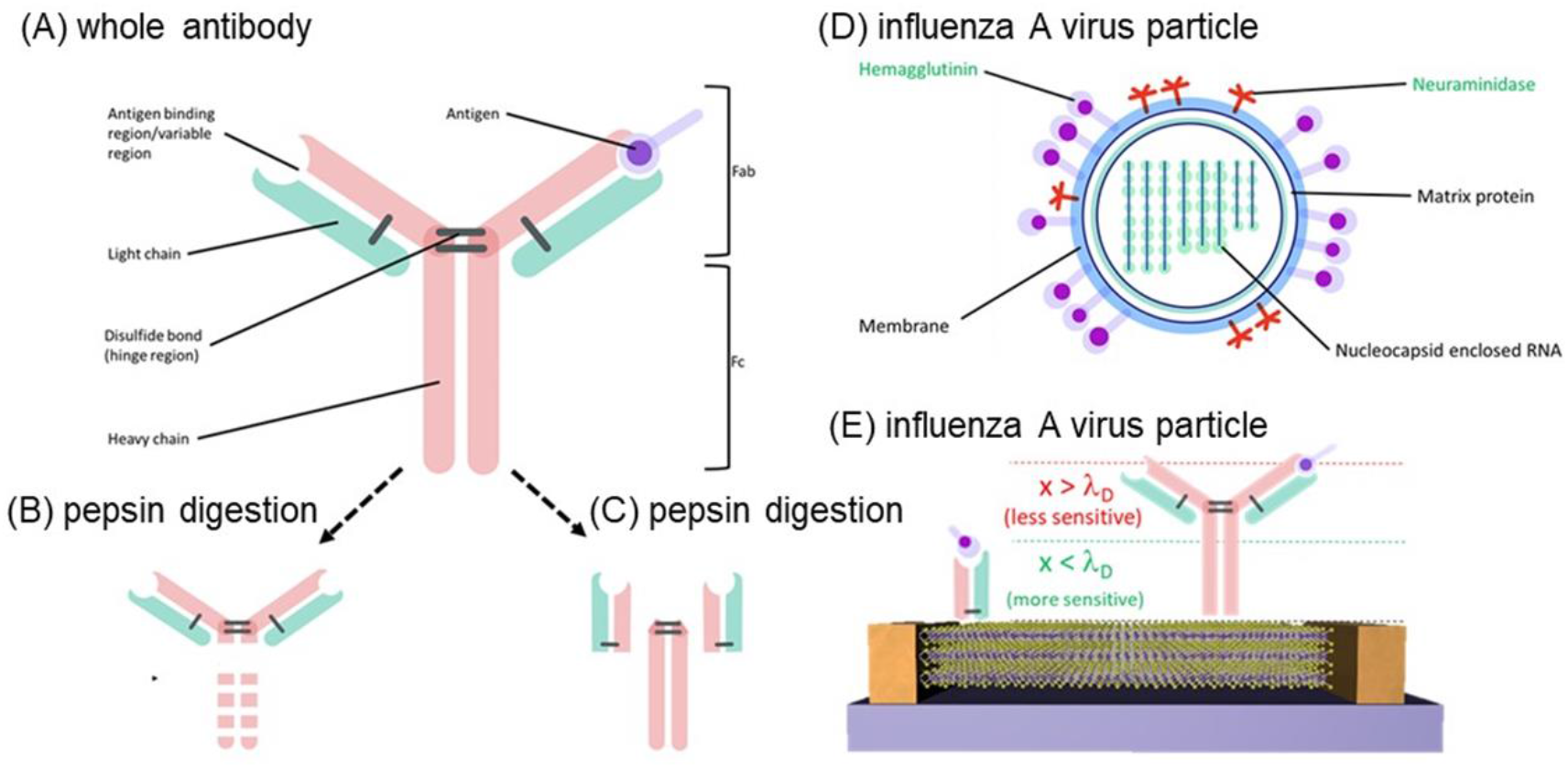
Antibodies demonstrate specific binding to a particular antigen with unique compositions and structures in their binding domains, however the general structure shown in (A) for all antibodies is the same. Antibody cleavage in the desired location (B-C) can be attained using different protease treatments. Virus particles have unique structures as well. A schematic of influenza A (D) is shown with potential target antigens for sensing highlighted in green. Binding of hemagglutinin or HA glycoprotein on the surface of the virus in (D) in the antigen binding region of the antibody is shown in (A, upper right). Fragments of the antibody generated via papain digestion are expected to bind to sulfur surfaces defective with sulfur vacancies (E) in a favorable orientation for interactions with the sample and also bring the antigen closer to the sensor transducer surfaces, which enhances sensitivity if that distance (x) is on the order of a Debye length (λ_D_). Figures A-D used with permission from American Institute of Physics, C. Muratore and M.K. Muratore, *J. Vac. Sci. Technol. A* 38 (2020) 050804.

Utilization of the dual specific binding attributes of antibody fragments to transducer and antigen is an attractive means to achieve sensor selectivity. Antibodies are modular, soluble, Y-shaped molecules with three equally sized portions (Fig. 1A). There is a constant region that binds to interfaces with effector cells (various types of cells that bring about changes in the body, i.e., immune responses) and two variable regions that bind to either a specific sequence (linear epitope) or to a specific structure (non-linear epitope) comprising the antigen. Every antibody consists of heavy chains and light chains linked in a hinge region by disulfide bonds^2^. This structure allows for functional fragmentation by proteases. Examples include pepsin digestion resulting in two fragments (Fig. 1B) or papain digestion resulting in three fragments (Fig. 1C), two of which are designated as Fab fragments interacting with proteins comprising the virus. In the case of influenza A (Fig. 1D), hemagglutinin (HA) and neuraminidase are glycoproteins known to interact with receptor binding domains of antibodies^23^ as shown in Fig. 1E. The other fragment, the part that interacts with the effector cells or molecules is called the Fc fragment (fragment crystallizable). In this work, laser-crystallized hexagonal MoS_2_ transducers were functionalized with monoclonal Fab fragments (Fig. 1E) for selective binding to SARS-CoV-2 or Influenza A glycoproteins. Additional pathogens may be detected with the same devices by attaching any complementary antibody fragment (such as the CR3022 Fab in Fig 2A) to the photonically-crystallized semiconducting MoS_2_ surface which intrinsically exhibits sulfur defects for bonding of the fragment in a conformation orienting its antigen binding sites towards the sample for analysis. Another bio-macromolecule known to have an affinity for the trimeric SARS-CoV-2 spike glycoprotein is the angiotensin converting enzyme 2 (ACE2) protein domain^24-25^ (Fig. 2B), which was also examined as a bio-recognition element (BRE) in the current work. The protein was terminated with histidine, reported to bind to MoS_2_ ^26-27^ such that its binding domain was expected to possess an orientation toward the sample.

**Figure 2:**
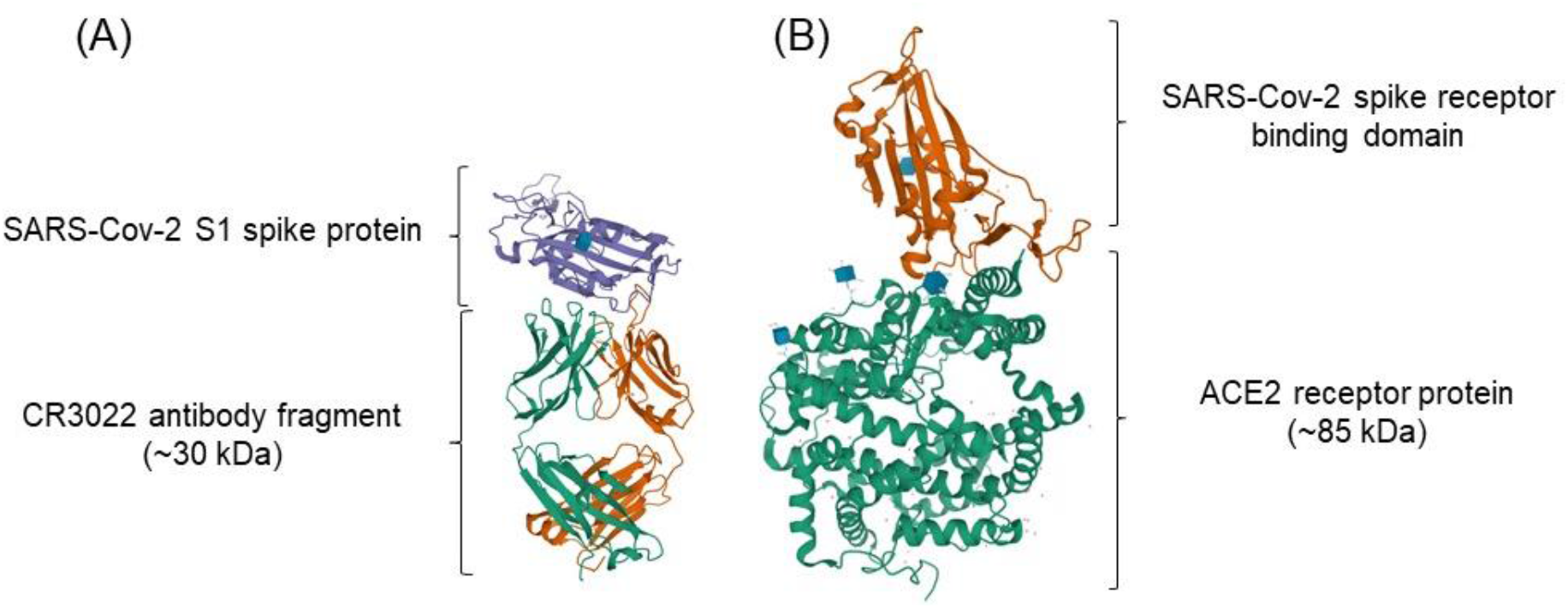
Crystal structures of COVID-19 antigens with complimentary receptors (a) CR3022 antibody Fab fragment^32^ and (b) human ACE2 receptor^33^. The sequence for (b) is found as Supporting Information.

Antibody fragment functionalization and attachment of ACE2 protein fragments to 2D MoS_2_ transducer surfaces was investigated as a means to produce multiplex electronic sensors for detection of COVID-19 and Influenza A antigens in liquid samples resembling saliva with picogram/mL detection on a single, compact, disposable electronic chip device with nine individual sensor transducers. Fabrication involved existing industrial processes (e-beam evaporation, sputtering, and laser-writing) with a low estimated manufacturing cost per chip. Each of the nine transducers on the chip may be functionalized with the BRE(s) for pathogen(s) of interest in the adjacent saliva sample. Saliva was selected as a target test fluid for the sensors not only because it is more easily accessible than a sample from the nasopharynx (currently used for most institutional COVID-19 tests), but is also reported to contain a higher SARS-CoV-2 viral load^28^. While the sensors presented in the current work are for selective detection of SARS-CoV-2 and Influenza A, the devices are modular, meaning that the general structure of all human antibodies can be utilized to functionalize sensor transducers for selective detection of diverse pathogens. This feature is attractive for meeting sudden worldwide needs as chips with the underlying sensor device structure can be stockpiled and later functionalized on demand with antibodies, antibody fragments, or other BREs for pathogens of interest. Ultimately, proteins from the receptor binding domain of the antibody rather than the entire antibody fragment may be used to reduce cost and increase sensitivity^29^ as well as the ease with which the sensor could be fabricated, stored, and used as complex biomolecular assemblies such as antibodies and even fragments are susceptible to denaturation or other undesirable changes in conformation under ambient conditions.

Influenza A H5 Monoclonal Antibody fragments prepared via papain digestion were obtained (Virusys IA247) with a reported concentration of 470 μg/mL. The complementary hemagglutinin (HA) antigen was obtained from Abcam (217659). Whole SARS-CoV-2 spike protein antibodies known to bind to the S1 spike protein antigen^30^ (CR 3022, Abcam 273073) were digested using a Pierce Fab Preparation Kit (ThermoFisher 44985). SDS PAGE was performed on a mini-PROTEAN system (Biorad) to analyze protein digestion. The digested proteins were loaded into a precast 20% gradient polyacrylamide gel (Biorad) using 2x Laemmli sample buffer alongside a Precision Plus Protein Kaleidoscope ladder (Biorad 1610375) and run with a 25 mM Tris, 192 mM glycine, 0.1% SDS, 8.3 pH running buffer. An EZBlue Gel stain (Sigma-Aldrich G1041) was used to visualize protein separation in the gel and analyze results of the digestion (see Fig. S1). A nanophotometer (Implen, Denville Scientific) was used to determine the CR3022 Fab fragment concentration estimated at 158 μg/mL buffered in 7.4 pH phosphate buffer solution. The complementary antigen was the SARS-CoV-2 S1 spike protein (Abcam 272105). The histidine-terminated human ACE2 RBD protein fragment (Abcam 151852) at 700 ng/mL in PBS buffer at 7.4 pH was applied and tested with the complementary SARS-CoV-2 spike glycoprotein peptide (Abcam 273063) and also the same S1 spike protein (Abcam 272105) used for testing antibody fragment functionalized sensors. For all tests, antibody fragments or proteins were applied to the sensor surface at their maximum concentration. Influenza A and all SARS-CoV-2 antigens were buffered with PBS at a pH of 7.4 to match the typical higher pH of extracted saliva^31^.

A schematic of the devices and their fabrication method is shown in Fig. 3. Metal contacts (15 nm Cr/35 nm Au) with a gap of 85 μm were applied via electron beam evaporation to shadow-masked glass substrates (Corning Eagle XG of 500 μm thickness). The substrates and contacts were coated with continuous a thin layer (3 nm) of amorphous MoS_2_ via room-temperature magnetron sputtering as described in detail elsewhere^34^ (AFM results shown in Fig. S2). The amorphous material is then converted into 2H-MoS_2_ by focusing the output of a continuous-wave, argon-ion laser (Coherent Innova 90C) with a central wavelength of 514 nm was focused through a 100x objective lens onto the amorphous MoS_2_ surface with a 1/e^2^ diameter of approximately 20 μm at a power of 125 mW^34-36^. The beam was then rastered over a 190 x 85 μm area of the surface at a speed of 100 μm/s, while the sample was contained in ultra high purity argon at 11 Torr to selectively convert the amorphous MoS_2_ to nanocrystalline hexagonal MoS_2_ (2H-MoS_2_) as verified by Raman spectroscopy. The spacing between individual laser lines rastered over the surface was 6 μm, resulting in a uniform region of 2H-MoS_2_. Laser writing was also used to isolate each device by ablating a line of material between the devices with different geometries to tune the resistance across the device from 120-1200 kΩ.

**Figure 3:**
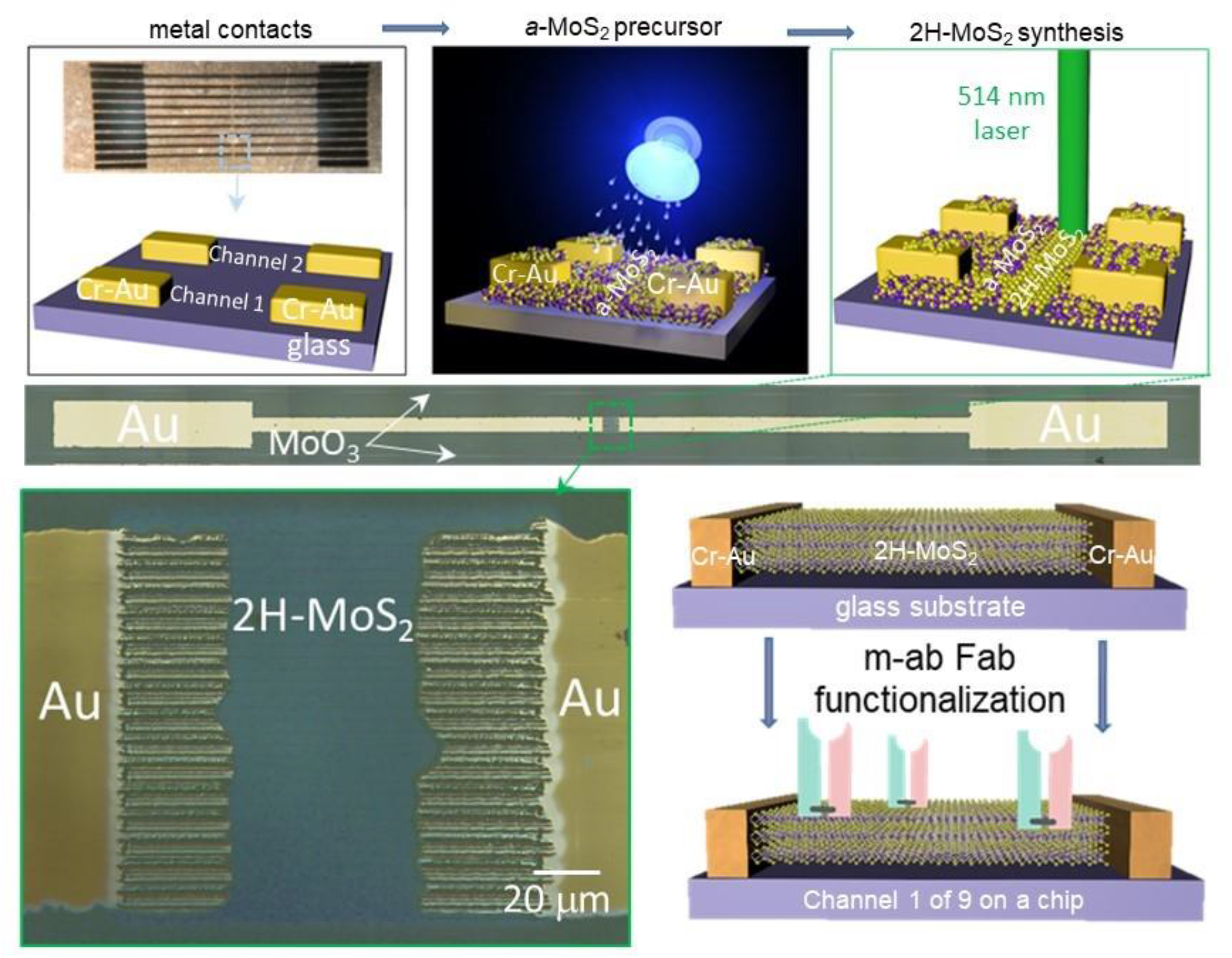
Schematic of sensor fabrication steps (top) and sensor geometry (bottom) with optical images of a complete single sensor on a chip (middle) and of the laser-annealed area comprising the sensor transducer (bottom left). A schematic of one of the nine devices on the chip is shown bottom right.

To measure the sensor response, the two-point resistance of the as-fabricated sensors was measured with a Signatone 1160 probe station driven by an Agilent 4156C Semiconductor Analyzer. Two-point resistance measurements were made by sweeping the current from 1 pA to 100 nA and measuring the voltage between contacts. The measured voltage never exceeded 500 mV to avoid measurement-induced electrochemical reactions at the sensor surface (e.g. water hydrolysis, tyrosine oxidation (0.7/1.4V depending on protonation state). The initial resistance between contacts was measured with a fixed probe spacing of 15 mm (example measurements shown in S3). Immediately after the resistance measurement, a 1 μL drop of BRE in solution was pipetted *in situ* onto the sensor area between the contacts using the probe station microscope. An initial resistance measurement was recorded 5 seconds after BRE application, and additional measurements were made over a period of three minutes. A 1μL drop of buffered antigen was then applied to the sensor region and resistance measurements were made in 5-60 second intervals thereafter. The electrical resistance of the buffered antibody and antigen solutions is 2-3 orders of magnitude higher than the resistance of the active sensor material (depending on the initial resistance of the sensor, R_o_); therefore it is presumed that electrical current was primarily conducted through the MoS_2_ sensor material.

After functionalization with 470 μg/mL (with expected surface coverage of approximately 10^9^ cm^-2^), devices with R_o_ = 120 kΩ were exposed to HA antigen concentrations from 2-2000 pg/mL. The response with time for application of 10^−12^ g/mL antigen was immediate as shown in Fig. 4b with the first measurement with applied antigen accumulated 5 seconds after introducing the antigen to the sensor. The decrease in resistance upon addition of both antibody and antigen is consistent with binding of negatively charged species to the p-type 2H-MoS_2_ semiconducting material^37^. Based on the 7.4 pH used for the measurements here and the reported isoelectric point for HA of 6.5-7^1^ it was anticipated that the antibody fragments and the antigen would both possess a net negative charge. Sensors functionalized for detection of influenza A were also exposed to the SARS-CoV-2 S1 spike protein. While a change in resistance was observed (Fig. 4a, red data points), its magnitude was significantly less than that measured for identical concentrations of the proteins complementary to the antibody, and the response did not vary with antigen concentration thereby indicating selective HA antigen binding.

**Figure 4:**
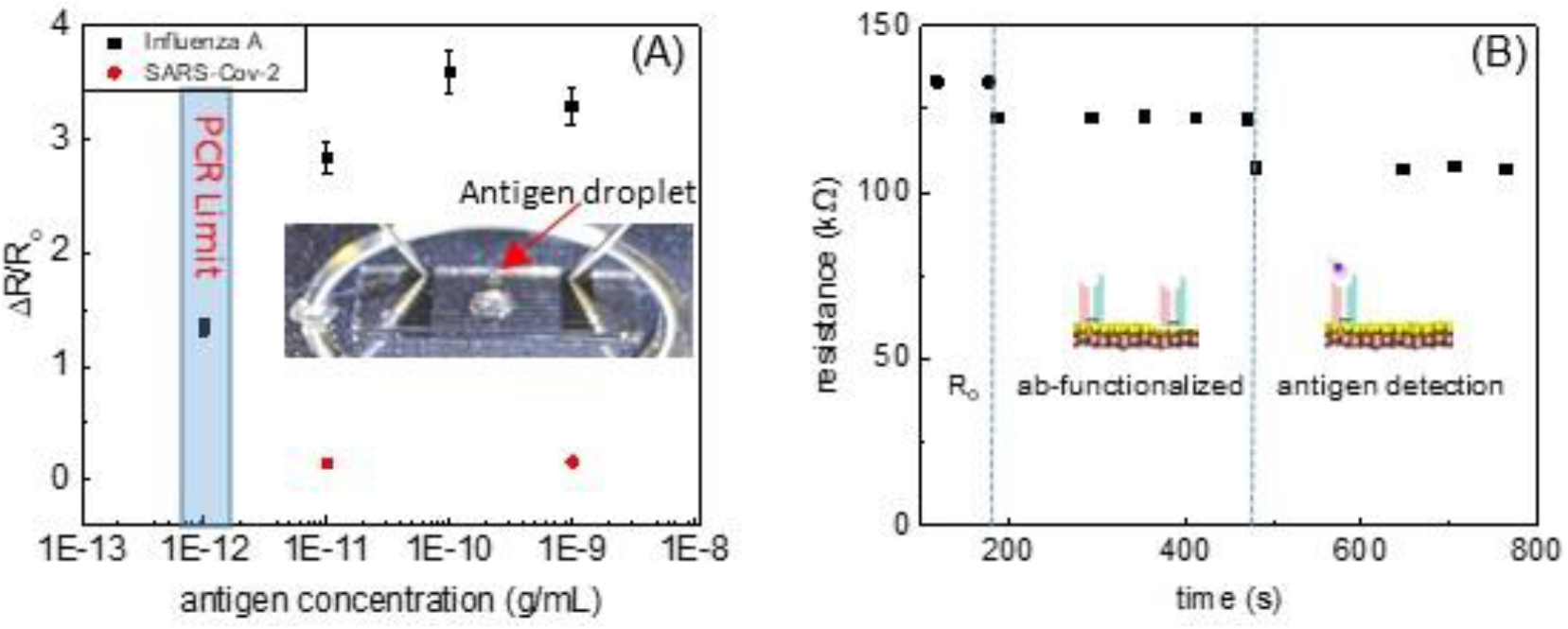
Sensor measurement results for (a) influenza A antibody Fab fragment functionalized sensor subjected to HA and SARS-CoV-2 spike proteins. Inset shows a photograph of the sensor during the measurement. The change in resistance with time (b) for antibody functionalization and after exposure to 1 pg/mL HA. The PCR limit is indicated as pg/mL (∼100,000 antigen proteins/mL) assuming one copy detection with ∼100 spike proteins per copy for PCR.

For the SARS-CoV-2 measurements, sensor devices with a range of initial resistances R_o_ = 120-1200 kΩ were investigated. The initial device resistance was related to the total area of the transducer, which was isolated by laser writing MoO_3_ around the device (as shown in Fig.3). The amorphous MoS_2_ precursor material will also respond to functionalization and application of antigen, but with less sensitivity than the crystallized hexagonal MoS_2_ (*h*-MoS_2_), therefore the larger resistance is indicative of a larger effective sensor as the 1 μL drops of applied BRE possessed an area larger than the 150 μm x 150 μm transducer area. Devices with enlarged sensor areas (i.e., devices with higher values of R_o_) resulted in higher sensitivity as shown in Fig. 5A. The sensors with the largest resistance (R_o_ = 1200 kW) show a response to pg/mL antigen concentrations.

**Figure 5:**
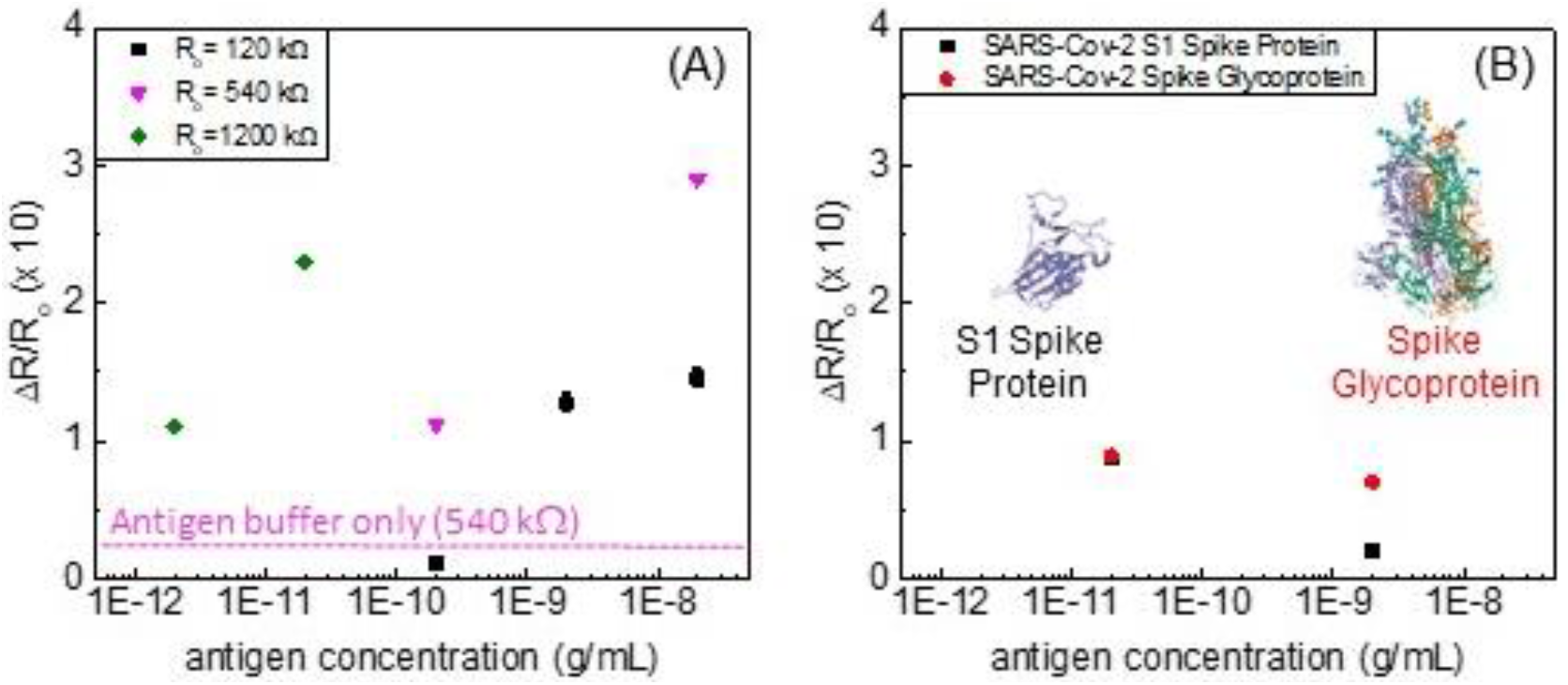
The sensors tested in (A) show responses to SARS-CoV-2 S1 spike protein vary with initial resistances (R_o_), with pg/mL antigen detection for sensors with higher R_o_. The response of a sensor exposed to PBS buffer with sodium azide at the concentration matching that of the antigen sample at 10^−10^ pg/mL was applied to a sensor with R_o_ = 540 kW. In (B), a small response to two different SARS-CoV-2 spike protein antigens with no dependence on concentration is shown for R_o_ = 1200 kΩ sensors functionalized with the ACE2 protein.

The resistance changes for the sensor functionalized with human ACE2 receptor protein are shown in Fig. 5B. The resistance decreased slightly upon functionalization with the protein and decreased further upon introduction of antigen, however the decrease did not vary with antigen concentration and was smaller than that observed for antibody fragment functionalization. The lack of responsivity is likely due to the increased separation of antigen from antibody for the larger BRE. Computational explorations including shorter sequences that maintain the key binding epitope of other proteins and peptides binding the SARS-CoV-2 spike protein are underway^38^ and will likely motivate subsequent studies of alternative BREs.

Characterization of a laser-written MoS_2_ transducer via Raman spectroscopy and x-ray photoelectron spectroscopy before BRE/antigen exposure and after sensor testing and rinsing with PBS suggests strong binding of the antibody/antigen pairs to the surface. Comparison of Raman spectra (Fig. 6A) reveals a strong response in the MoS_2_ A_1g_ peak (408 cm^-1^), consistent with reports that this peak is more sensitive than the E_2g_ (383 cm^-1^) peak to changes in phonon-hole coupling induced by doping^39^, and may explain both the broadening of the A_1g_ peak in addition to its reduction in intensity measured after testing. Sensors in the as-fabricated and post-test states were also investigated via x-ray photoelectron spectroscopy (XPS). A nitrogen peak (Fig. 6B) appears adjacent to the Mo 3p_3/2_ peak for the sensor after testing in comparison to the lone Mo peak for the as-fabricated transducer analyzed before sensor testing. The sulfur spectrum, also measured via XPS, is shown in Fig. 6C. There appears to be a subtle difference in the shape of the S 2p spectra for the sensors before and after testing, however applying the fitted peaks for the sulfur doublet for the as-fabricated device results in a fit that matches the peak shape of the as-measured spectrum for the sample after testing. While the presence of sulfur vacancies in the laser-written MoS_2_ receptor was confirmed via resonant Raman spectroscopy (S4), detection of sulfur termination of the as-fabricated MoS_2_ receptor surface with the antibody fragments may be suggested by the XPS data^40-41^, but the signal to noise-ratio was too low for a conclusive measurement. Additional high-resolution XPS spectra appear in SI5, in addition to more details regarding fitting of the spectra shown in Fig. 6.

**Figure 6:**
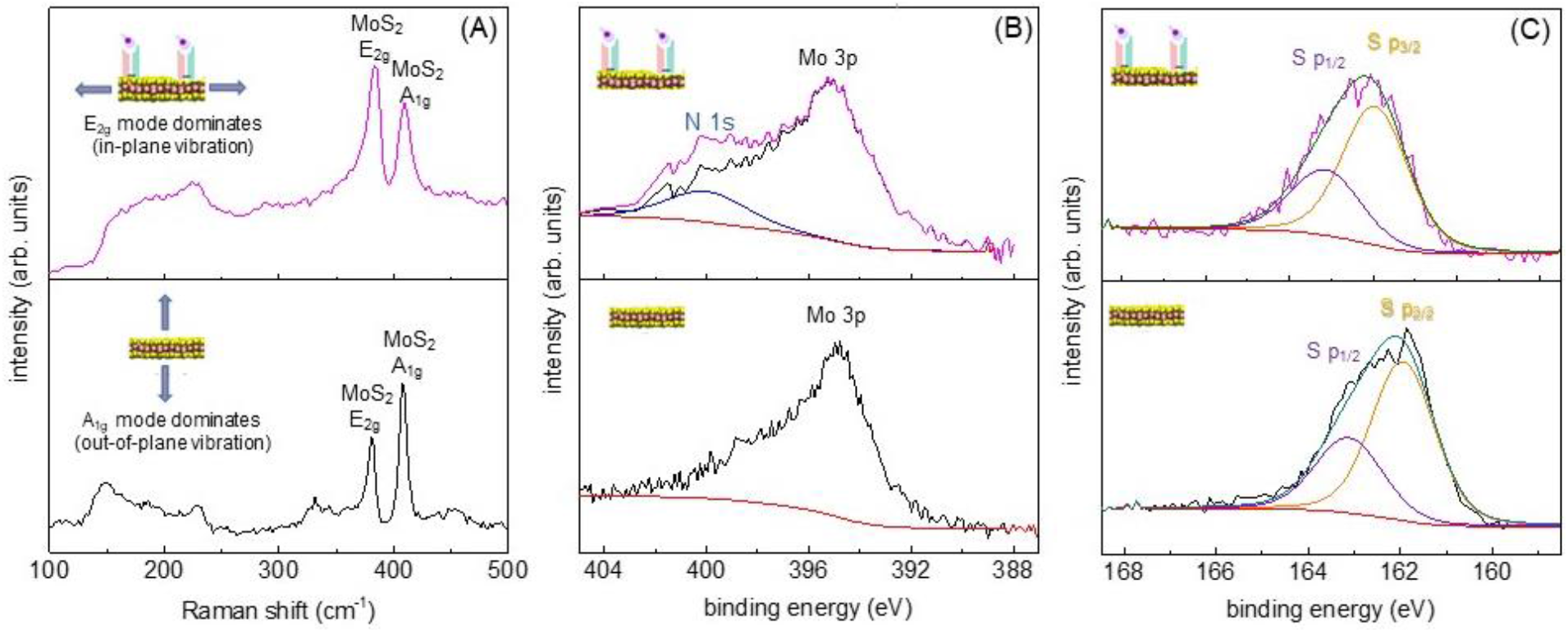
Materials characterization shows effects of biomolecular binding on influenza A sensors before (bottom) and after (top) testing with (A) Raman spectra showing preferential suppression of out-of-plane (A_1g_) vibrational modes for MoS_2_ transducer materials after testing indicative of electronic doping. High-resolution XPS spectra of (B) nitrogen peaks appearing adjacent to the molybdenum 3p peak after functionalization and indicating antibody/antigen binding, and (C) sulfur XPS peaks before and after sensor testing.

In conclusion, electronic antigen sensors demonstrating selective pg/mL virus detection for Influenza A and SARS-CoV-2, showing sensitivity competitive with institutional state-of-the-art tests, were fabricated via a scalable, economical, high-throughput fabrication approach integrating inexpensive, naturally abundant materials such as recyclable glass and small volumes of the mineral molybdenite (MoS_2_) to reduce impact of the used sensor waste stream. The sensor yield from this fabrication approach was 100 percent, that is, all devices tested were functional. Differentiation between viruses at the pg/mL sensitivity level was demonstrated via bio-functionalizing the surface with antibody fragments generated by papain digestion of whole antibodies for influenza A and SARS-CoV-2 antigens. Results suggest antibody fragments were chemisorbed to the MoS_2_ surface and retained the ability to bind antigens. Biofunctionalization of the sensor surface with human ACE2 receptor protein did not enable recognition of SARS-CoV-2 spike glycoprotein binding events, likely due to its larger displacement of the antigen from the transducer surface. It is anticipated that binding of antibody fragments to a MoS_2_ surface for a device with an analogous sensor geometry will yield similar sensitivity for any pathogen with a known complementary antibody, defining the sensors as truly modular in that the devices can be adapted on demand to meet a timely diagnostic need. Investigations of alternative BREs may result in even more affordable and robust devices.

## Data Availability

All data is available upon request

## Author Contributions

The manuscript was written through the contribution of all authors. All authors have approved the final version of the manuscript.

